# A repeat pattern of founder events for SARS-CoV-2 variants in Alaska

**DOI:** 10.1101/2022.05.25.22275610

**Authors:** Tracie J. Haan, Lisa K. Smith, Stephanie DeRonde, Elva House, Jacob Zidek, Diana Puhak, Logan Mullen, Matthew Redlinger, Jayme Parker, Brian M. Barnes, Jason L. Burkhead, Cindy Knall, Eric Bortz, Jack Chen, Devin M. Drown

## Abstract

Alaska is a unique US state because of its large size, geographically disparate population density, and physical distance from the contiguous United States. Here, we describe a pattern of SARS-CoV-2 variant emergence across Alaska reflective of these differences. Using genomic data, we found that in Alaska the Omicron sublineage BA.2.3 overtook BA.1.1 by the week of 2022-02-27, reaching 48.5% of sequenced cases. On the contrary in the contiguous United States, BA.1.1 dominated cases for longer, eventually being displaced by BA.2 sublineages other than BA.2.3. BA.2.3 only reached a prevalence of 10.9% in the contiguous United States. Using phylogenetics, we found evidence of potential origins of the two major clades of BA.2.3 in Alaska and with logistic regression estimated how it emerged and spread throughout the state. The combined evidence is suggestive of founder events in Alaska and is reflective of how Alaska’s unique dynamics influence the emergence of SARS-CoV-2 variants.

## Introduction

Throughout the coronavirus disease 2019 (COVID-19) pandemic, variants of severe acute respiratory syndrome coronavirus 2 (SARS-CoV-2) have repeatedly emerged and spread, often circulating globally over a relatively short timeframe (Mullen et al., 2020). These variants frequently underwent mutations affecting viral phenotypes, such as increased transmissibility or immune escape, which contributed to epidemic waves of cases and hospitalizations occurring asynchronously across different regions at varying severities (Alcantara et al., 2022). The sequential wave dynamics of COVID-19 have likely been influenced by a multitude of epidemiological factors, such as host immunity and vaccination coverage, social measures aimed at suppressing spread, and the viral characteristics, including transmissibility and the moderately higher mutation rate for an RNA virus (Saha et al., 2021; Kirby, 2021; Tao, 2021).

For many regions of the world, the first notable COVID-19 epidemic wave attributed to a variant occurred near the end of 2020 into early 2021. During this wave, the Alpha variant (lineage B.1.1.7; Rambaut et al., 2020), which showed evidence of increased transmissibility, became the most prevalent variant for most places globally (Volz et al., 2021). Unlike other regions, including within the contiguous 48 states of the United States (hereafter referred to as Lower 48), Alaska’s dominant lineage was B.1.1.519 throughout early 2021, which was similar to Mexico in late 2020 (Haan et al., 2022; Rodríguez-Maldonado et al., 2021). The timeline in which Alpha and B.1.1.519 emerged in Alaska paired with the striking difference in prevalence between Alaska, which had a peak B.1.1.519 prevalence of 77.9%, and the Lower 48, which had a peak prevalence of 4.9%, was indicative of a B.1.1.519 founder event in Alaska (Haan et al., 2022). Since this initial deviation from the Lower 48, Alaska has displayed similar patterns of variant emergence and spread both with the sweep of the Delta variant in early August 2021 followed by the sweep of Omicron beginning in December 2021 (CDC, 2021).

Although the recent emergence of Omicron in Alaska was initially similar to that of the Lower 48, the emergence of sublineages within larger variant classifications has been distinct. These Omicron sublineages have shown a great degree of divergence that has led to concerns about antibody evasion and the possibility of repeat infections of Omicron sublineages (Stegger et al., 2022). Mutations include a spike protein (S) R346K alteration in BA.1.1 (also known as B.1.1.529.1.1) and 8 unique S mutations in BA.2, which lacks 13 S alterations found in BA.1. The Omicron sublineage reported in this study, BA.2.3, encodes several key amino acid changes including A2909V in ORF1a and L140F in ORF3a. These changes may have provided growth advantages such as antibody evasion and increased reproductive rate that allowed Omicron lineages to displace Delta initially (Iketani et al., 2022; Smith et al., 2022).

Here, we use genomic data readily available via the Global Initiative on Sharing All Influenza Data (GISAID) to describe the pattern of emergence and spread of the Omicron sublineage BA.2.3 (also known as B.1.1.529.2.3) in Alaska. We contrast this pattern with observations in the Lower 48 and several major US states including California, New York, and Washington. Using data generated through genomic surveillance efforts, we explore the pattern in Omicron sublineages of Alaska similar to that of the founder event that occurred with B.1.1.519 in early 2021 (Haan et al., 2022). These repetitive patterns of variant emergence are suggestive of repeat founder events in Alaska.

## Materials and Methods

### Retrieving and Analyzing SARS-CoV-2 Sequence Data for Alaska

On May 3rd, 2022, we downloaded 11,971 sequences from Alaskan samples available on GISAID for subsequent analysis (Shu & McCauley, 2017; Elbe & Merret, 2017). This readily available genomic data was in part generated by the *Alaska SARS-CoV-2 Sequencing Consortium*. The Consortium is a partnership between the University of Alaska and the Alaska Division of Public Health (AKDPH) with the aim to increase genomic surveillance of SARS-CoV-2 variants. Genome sequencing in Alaska is from a non-targeted sample of cases, which is the best available approximation of random samples despite potential disparate coverage across Alaska’s economic regions. We subset these data to the study date of 2021-11-29 through 2022-04-02 for a total of 3,132 genomes to estimate the prevalence of lineages per week on dates beginning on Sunday of each week.

Lineages were determined by running sequences through PANGO v1.8, Pangolin v4.0.6, and pangoLEARN v1.2.133, and Scorpio v0.3.17 (O’Toole et al., 2021). We estimated the prevalence of genomes in Alaska from the date of Omicron’s (B.1.1.529) first detection in Alaska on 2021-11-28 through 2022-04-03. All AY sublineages are aggregated into the group B.1.617.2 (Delta). All BA sublineages of Omicron except BA.1.1, BA.2, BA.2.3, and their sublineages are aggregated into B.1.1.529 (Omicron). Genomes that did not fall into these lineages were grouped together into the category ‘Not Emerging Lineage.’

### SARS-CoV-2 Sequence Data for the Lower 48

On April 12th, 2022, we downloaded metadata, including lineage assignment using the same version of Pangolin (v4.0.6) for all sequences available on GISAID and filtered for sequences from the United States of America (USA) collected between 2021-11-28 to 2022-04-12. We removed cases from Alaska, Hawaii, and US territories to limit our comparisons to the lower 48 contiguous states. We also analyzed data at a state level for New York, California, and Washington. GISAID metadata was used to calculate the prevalence of variants, which was an approximation based on percent of sequenced cases each day.

### Visualizations, statistical analyses, and Nextstrain build

We generated visualizations in RStudio (v 1.4.1106) using packages ggplot2 (v 3.3.5), ggpubr (v 0.4.0), tidyverse (v 1.3.1), and lubridate (v 1.7.10). A generalized linear model using the logit link function with the base R stats (v 3.6.2) glm tool was used to generate estimates of prevalence of BA.2.3 over time for Alaska and two economic regions; the Anchorage-Mat Su and Gulf Coast. For these models, the daily percent of sequenced cases assigned to BA.2.3 (i.e., daily prevalence of BA.2.3) was used as the dependent variable with time from 2022-01-01 through 2022-04-02 used as the independent variable. Regressions were plotted using geom_smooth in ggplot2. We generated a Nextstrain (cli v-3.2.4) build to examine the phylogenetic relationship of BA.2.3 in Alaska compared to global sequences. We generated this tree using GISAID’s Global Nextregions for context, all BA.2.3 cases from Alaska, and global cases of BA.2.3 from 2021-12-02 through 2022-02-06 that included all cases from before 2022-01-01 and then downsampled to a fifth of the sequences randomly after that date. We colored tree tips by countries of significance including the Philippines, South Africa, Japan, India, and the USA split by ‘USA-Alaska’ and ‘USA-Other.’ Other country’s cases were masked from the tree visualization.

## Results and Discussion

### Higher prevalence of Omicron lineage BA.2.3 in Alaska versus the Lower 48

To examine how the emergence of BA.2.3 differs between Alaska and the Lower 48, we determined the date of first detection and prevalence of BA.2.3 over time in both locations. The first Alaska case assigned to Omicron was detected in the Anchorage-Mat Su region, the most populated region of Alaska, on 2021-11-28. Within four weeks of first detection (by the week of 2021-12-19), Omicron had outcompeted Delta in terms of prevalence both in Alaska and the Lower 48 (Figure 1). By the week of 2022-01-16, Delta was detected in less than 1% of sequenced cases for both Alaska and the Lower 48. Omicron cases during this week were dominated by the sublineage BA.1.1 in both Alaska, at 66.3% prevalence, and the Lower 48, at 67.2% prevalence (Figure 1). While BA.1.1 was dominant the week of 2022-01-16 in both locations, BA.2 and sublineages were just starting to be detected in the United States. By the week of 2022-01-16, in the Lower 48 BA.2 comprised only 0.2% of sequenced cases whereas in Alaska no cases of BA.2 had been detected. However, the sublineage BA.2.3 was found in 2.7% of sequenced cases in Alaska by this week whereas in the Lower 48 BA.2.3 represented only 0.1% of cases. By 2022-02-27, BA.2.3 comprised the majority of cases in Alaska (45.3%) compared to 6.1% in the Lower 48. At the same time, BA.2 comprised 9.4% of Lower 48 cases and only 2.5% of Alaska’s cases. Although by March BA.2.3 started increasing in prevalence in the Lower 48, BA.2 already displaced BA.1.1, the previously dominant lineage. By the last week of March, 2022, BA.2.3 comprised 74.3% of cases in Alaska and 19.2% of cases in the Lower 48. These stark differences in prevalence over time reflect the divergent patterns of emergence of BA.2.3 in Alaska versus the Lower 48.

**Figure 1.**
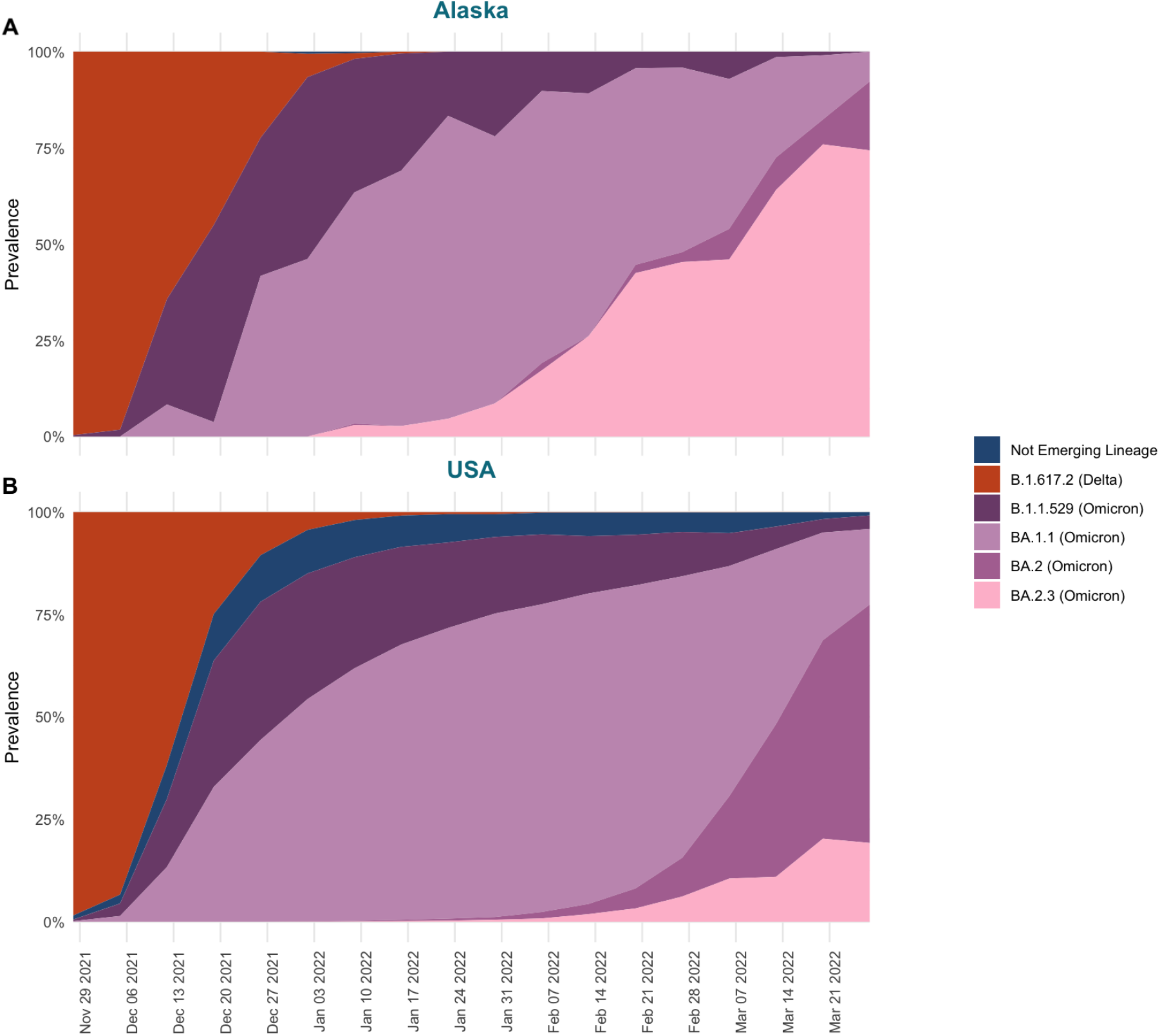
The percent of sequences by week (estimated prevalence) colored by SARS-CoV-2 lineages detected from 2021-11-28 to 2022-04-03 in (A) Alaska and (B) the Lower 48. BA lineages of Omicron except BA.1.1, BA.2, and BA.2.3 are aggregated into B.1.1.529. BA.2 includes all sublineages of BA.2 detected except BA.2.3.

Given the Lower 48 is an aggregate of many distinct, yet connected, communities, we examined the prevalence of sublineages at a finer geographic scale. This finer scale was at a state level for several populous US states including California, Washington, and New York. Each of these states reflected a similar pattern in BA sublineages as the overall Lower 48 with a low prevalence of BA.2.3 compared to Alaska over the study time period. The week of 2022-02-13 BA.2.3 already comprised 25.3% of sequenced cases in Alaska and only 1.8% in the Lower 48, 1.8% in New York, 2.2% in California, and 2.4% in Washington (Figure 2). The week of 2022-03-13 when BA.2.3 comprised a majority of the cases sequenced in Alaska at 67.9% whereas the other states had much lower and variable prevalence in the Lower 48 (10.9%), New York (5.5%), California (16.2%), and in Washington (17.6%) (Figure 2). By the last week of March, the only other state with a BA.2.3 prevalence greater than 40% was California.

**Figure 2.**
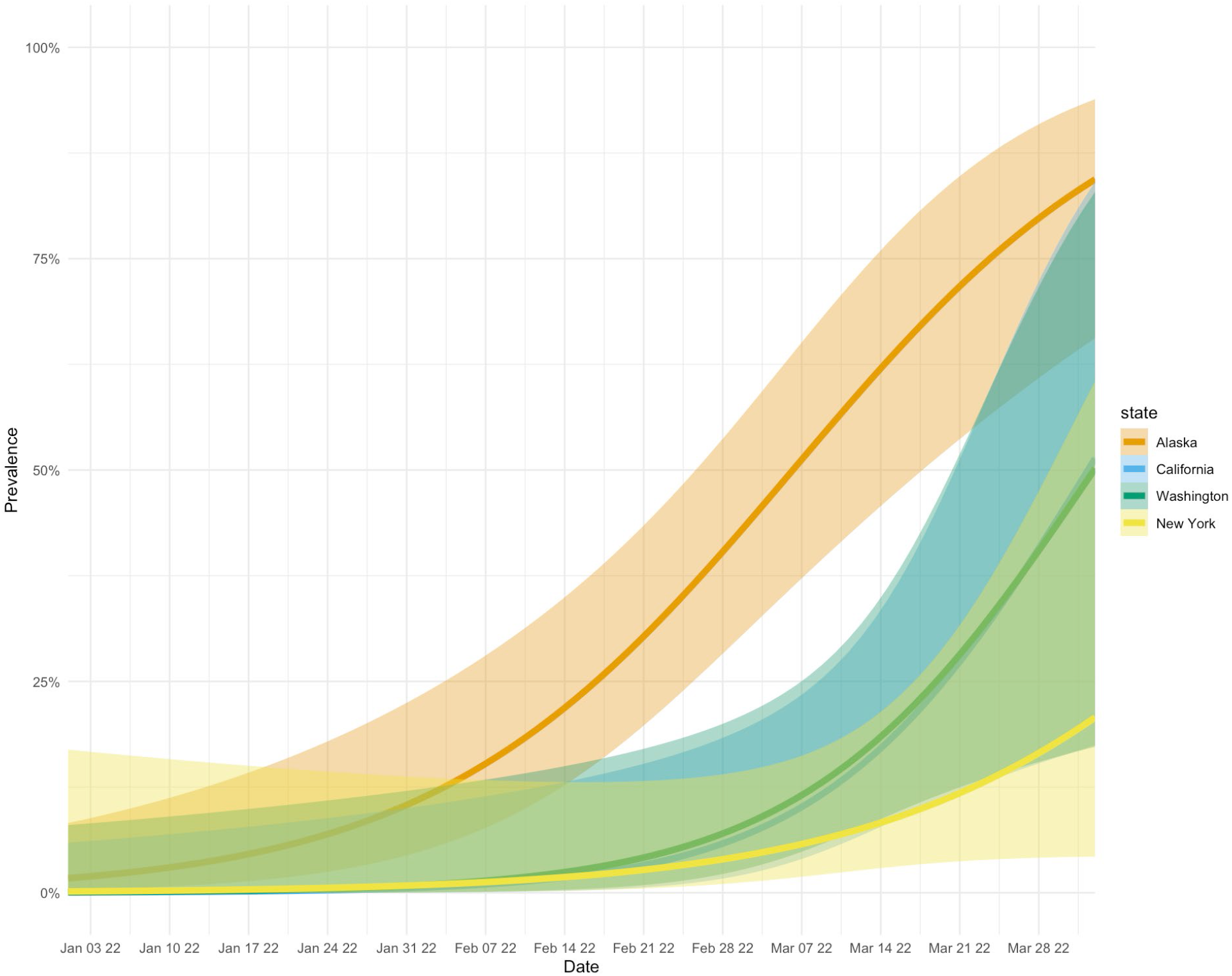
The percent of sequences by week (estimated prevalence) belonging to BA.2.3 colored by states including Alaska, California, New York, and Washington detected from 2021-11-28 to 2022-04-03.

Although selective advantages, such as transmission potential, posed by SARS-CoV-2 variants have played a key role in their emergence over the course of the pandemic, changes in variant prevalence can also be attributed to founder effects (Attwood et al., 2022). In the context of SARS-CoV-2, and other viral pathogens, founder effects result from a chance colonization event allowing a new population of viral lineages to emerge. When a chance colonization event occurs, the growth of a new population of viral lineages can give the impression that one lineage has a growth advantage over others (Rambaut et al., 2004; Ruan et al., 2021). The apparent difference in the emergence of BA.2.3 between Alaska, the Lower 48, and other US states highlights a potential founder effect in which BA.2.3 is acting as the founding sublineage in Alaska. It may have become dominant here and not in other locations because of the timing of emergence and social factors that rendered Alaska communities as a naïve population susceptible to infection by BA.2.3.

The composition of SARS-CoV-2 lineages within Alaska and the Lower 48, each containing distinct sets of mutations that define them, were distinct at the time of BA.2.3’s emergence in Alaska. The main difference in community composition was the presence of other BA.2 lineages in the Lower 48 and their absence in Alaska (Figure 1). The absence of other BA.2 lineages in Alaska could have allowed for the founding of BA.2.3 in the population, similar to how other lineages with specific mutations emerged and became dominant in other locations throughout the pandemic (Ozer et al., 2021; Hodcroft et al., 2021). For example, in the summer of 2020, the 20E lineage of SARS-CoV-2, which had no evidence of increased transmissibility, became the dominant lineage in Europe, likely driven by its founder event in the population paired with the increased connectivity across Europe from travel over the summer months (Hodcroft et al., 2021). It was also suggested the emergence of variant of concern (VOC) Alpha (B.1.1.7) and the associated mutations, like D614G, in part could have been driven by the founder effect (Ozer et al., 2021; Tang et al., 2021). This was suggested because of the inconclusive results over the positive selection of those mutations and coinciding timing with the nexus of dispersal from Asia to Europe associated with the D614G mutation (Grubaugh et al., 2020). Given the emergence of other lineages in the Lower 48 even though BA.2.3 was detected around the same time for many locations, Alaska’s emergence of BA.2.3, and B.1.1.519 earlier in the pandemic, implicates repeat occurrences of variant emergence influenced by the founder effect (Haan et al., 2022).

### Modeling shows variable emergence of BA.2.3 across Alaska

Spatiotemporal variation in the emergence and spread of SARS-CoV-2 lineages has been observed at broad geographic levels. The CDC has reported on these regional variations by dividing the United States into ten regions that show distinct communities based on genomic surveillance (CDC, 2022). Here, using genomic surveillance data from Alaska, we found within-state variation in the emergence of BA.2.3 between major economic regions of Alaska (Figure 2; Alaska Department of Labor and Workforce Development, 2021). In Alaska there are six economic regions defined by the Department of Labor and Workforce Development: the Anchorage-Mat Su, Interior, Gulf Coast, Southeast, Southwest, and Northern regions in order from highest to lowest population. BA.2.3 was first detected from two cases collected on 2022-01-11 in the Gulf Coast and the Anchorage-Mat Su regions of Alaska. The Gulf Coast is the third most populous region of Alaska and just south of the most populated region, the Anchorage-Mat Su. While many economic regions across Alaska are only connected by air or boat transportation, the Gulf Coast and Anchorage-Mat Su regions are broadly connected via Alaska’s road system. When examining a model estimate of BA.2.3 prevalence over time, we found that the Gulf Coast region had the earliest emergence. For the state as a whole, prevalence was estimated to be at greater than 5% the week of 2022-01-19 (Figure 3A), the Anchorage-Mat Su didn’t reach 5% until 2022-01-25 (Figure 3B), and the Gulf Coast was estimated to reach greater than 5% prevalence on 2022-01-03, which was before BA.2.3 was even detected (Figure 3C). Having the model indicate 5% prevalence before first detection suggests BA.2.3 could have been present in the Gulf Coast region of Alaska before sequencing captured a case of BA.2.3; however, model uncertainty, indicated by the shading, is also consistent with BA.2.3 being absent until the first actual case was detected. In the Gulf Coast, BA.2.3 was estimated to comprise the majority of cases by 2022-02-16, whereas for the state as a whole this didn’t occur until weeks later, 2022-03-07 and for the Anchorage-Mat Su region this didn’t occur until 2022-03-11.

**Figure 3.**
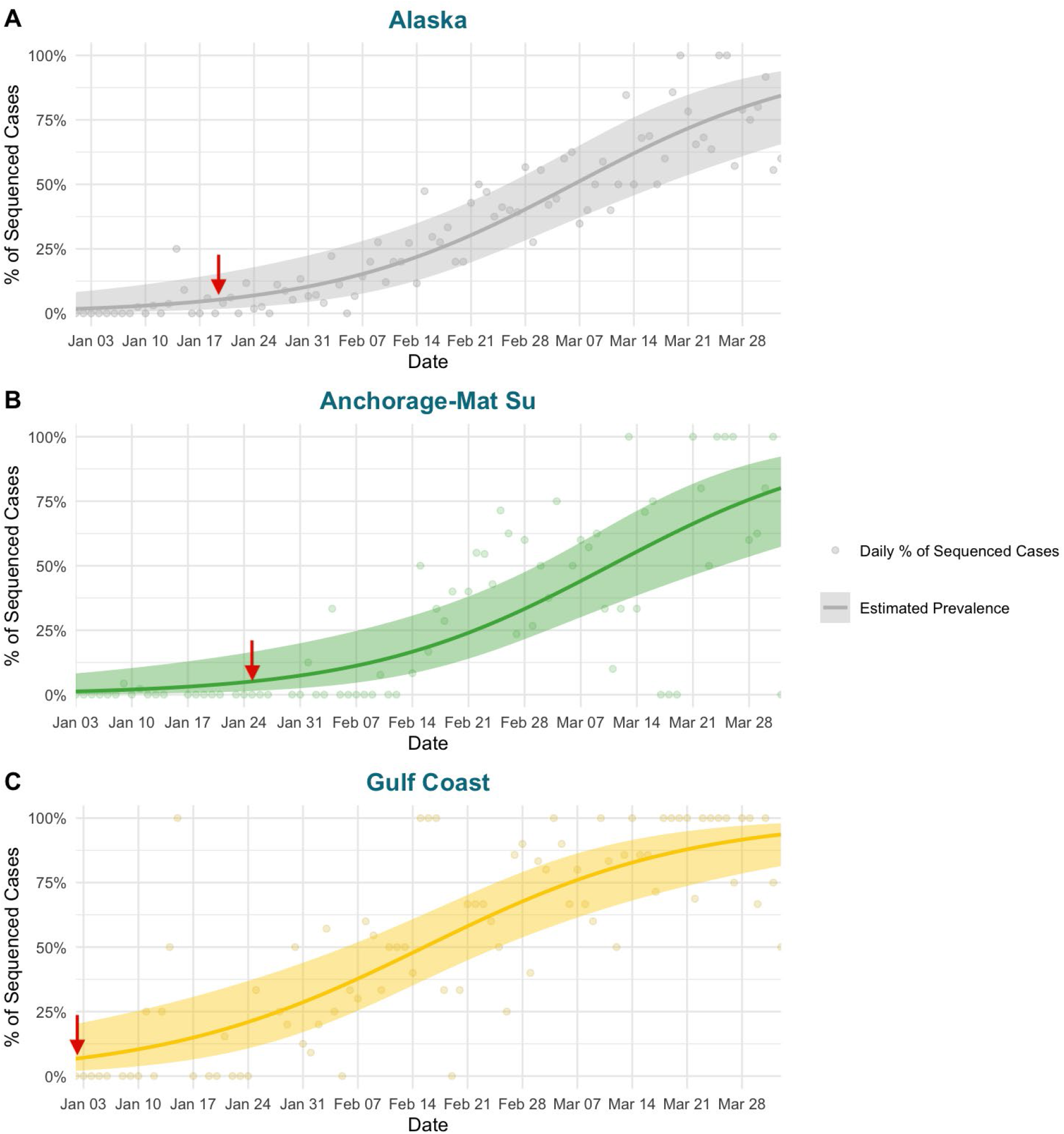
Logistic regression (line = regression; shaded region = standard error) estimating the prevalence of BA.2.3 over time in A) Alaska and the two economic regions of Alaska with a deep enough coverage of cases including (B) the Anchorage-Mat Su and (C) Gulf Coast. Points represent the daily percent of cases assigned to BA.2.3 used to calculate the regression. The red arrow highlights when the regression estimated BA.2.3 was at greater than 5% prevalence.

### Phylogenetics of BA.2.3 provides evidence of multiple introduction events

Based on global sequence data of BA.2.3 available on GISAID, we found that within Alaska there are two clades of BA.2.3 comprising the majority of Alaska’s cases. These two clades both appear to have emerged from cases originally detected in the Philippines, where BA.2.3 was first detected on 2021-12-02 (Figure 4). When considering cases by economic region of Alaska, there is no evidence that the two clades were introduced to each economic region independently given the interspersed nature of the cases (Figure 5). This suggests that there was mixing of cases between the two regions, rather than two separate introduction events. In other regions of the United States, BA.2.3 cases appear to have emerged from both the Philippines and a clade where South Africa and India cases appear to be dominant early in the tree. (Figure 4). By scaling the branch length by divergence, or number of mutations, we show how the majority of Alaska cases diverged from the early Philippines cases accumulating mutations with further spread in Alaska (Figure 4).

**Figure 4.**
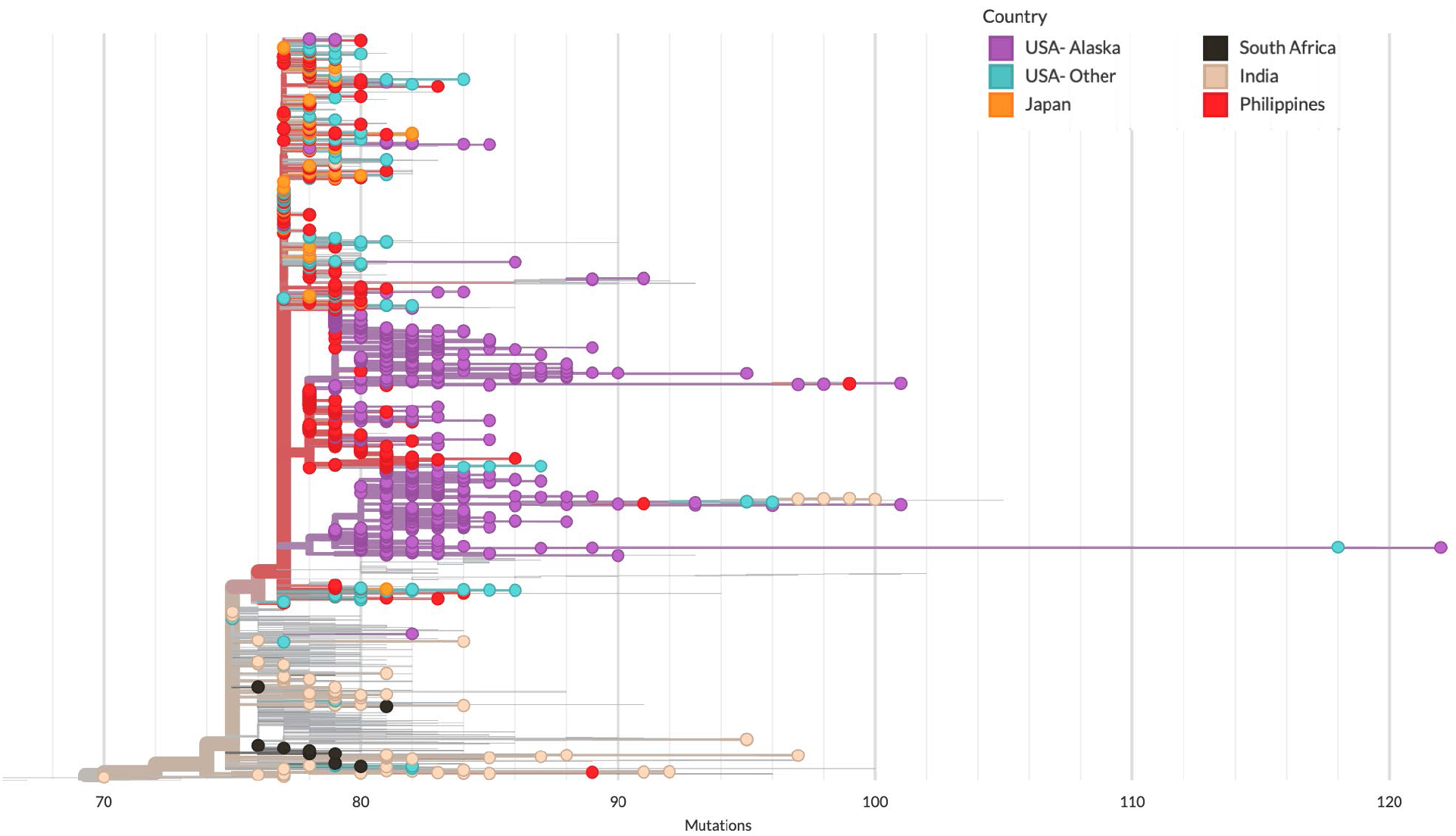
Phylogenetic tree of BA.2.3 cases with branch lengths represented by divergence of cases (number of mutations from Wuhan-Hu-1). Each point is a genome colored by country. Only countries that provide context for Alaska clades and clade origins are included in the visualization.

**Figure 5.**
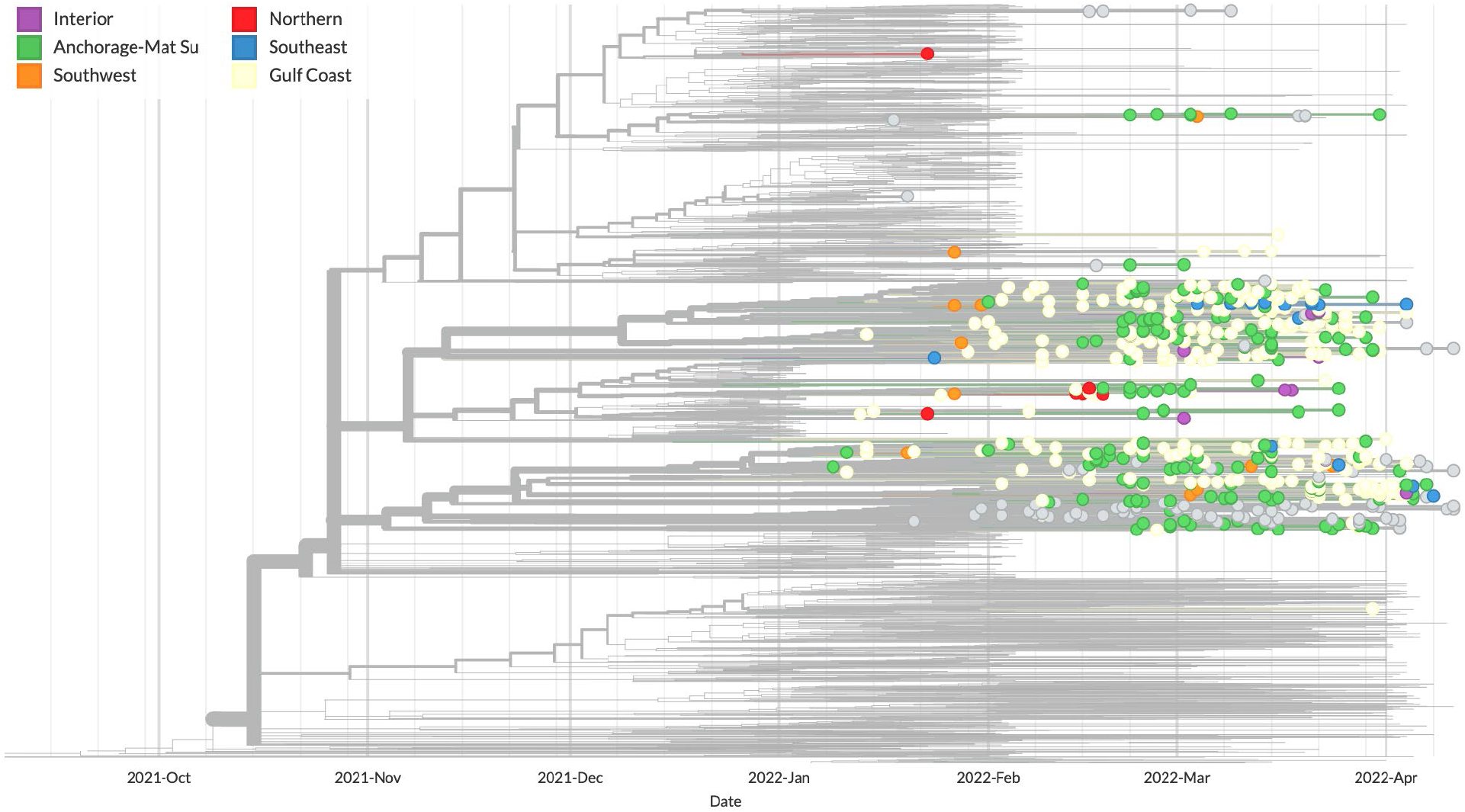
Phylogenetic tree of BA.2.3 cases with branch lengths represented by time. Tree includes all BA.2.3 cases from Alaska, all global BA.2.3 cases from December 2021, and all global cases after December 2021 through the first week of February 2022 downsampled for context. Only cases from Alaska are shown and are colored by economic region. Cases where the economic region is unknown are colored gray.

## Conclusions

Using genomic data available in the GISAID repository, we demonstrated the unique emergence and spread patterns of the SARS-CoV-2 sublineage of Omicron BA.2.3 in Alaska compared to the lower 48 contiguous states of the United States. Looking at a finer scale with several major US states, the same stark difference in prevalence of BA.2.3 was observed further highlighting the unique occurrence of BA.2.3 in Alaska. Our phylogenetic analysis paired with logistic regression revealed the potential ancestral origins of BA.2.3 cases in Alaska and how these clades within BA.2.3 emerged and spread by economic region of the state. These repetitive patterns of variant emergence with B.1.1.519 followed by BA.2.3 in Alaska are suggestive of repeat founder events which are reflective of how Alaska’s unique location influences the emergence of distinct SARS-CoV-2 variants

## Data Availability

All data used in this study are available online through the SARS-CoV-2 repository, global initiative on sharing all influenza data (GISAID). These findings are based on analysis of approximately 4,490 genomes accessible via EPI_SET_20220517as and 1,009,539 accessible via EPI_SET_20220517ge. Accession numbers for genomes of Alaska cases, metadata for the United States of America, and Global Nextregion data retrieved from the GISAID can be found using the EPI-SET identifiers at https://www.gisaid.org/

## Acknowledgments

We gratefully acknowledge all the researchers responsible for obtaining specimens and laboratories where genetic sequence data were generated and shared via the GISAID Initiative (https://www.gisaid.org), on which this research is based. Work presented here was supported by Epidemiology and Laboratory Capacity for Prevention and Control of Emerging Infectious Diseases (ELC) from the Centers for Disease Control and Prevention and a supplement award to Alaska INBRE, an Institutional Development Award (IDeA) from the National Institute of General Medical Sciences of the National Institutes of Health under grant number 2P20GM103395.

